# Consensus on COVID-19 vaccination in pediatric oncohematological patients, on behalf of infectious working group of Italian Association of Pediatric Hematology Oncology

**DOI:** 10.1101/2022.01.06.22268792

**Authors:** Simone Cesaro, Paola Muggeo, Daniele Zama, Monica Cellini, Katia Perruccio, Antonella Colombini, Francesca Carraro, Mariagrazia Petris, Valeria Petroni, Maurizio Mascarin, Francesco Baccelli, Elena Soncini, Rosamaria Mura, Milena Laspina, Nunzia Decembrino, Roberta Burnelli, Stefano Frenos, Elio Castagnola, Maura Faraci, Cristina Meazza, Federica Barzaghi, Maria Rosaria D’Amico, Maria Capasso, Elisabetta Calore, Ottavio Ziino, Angelica Barone, Francesca Compagno, Laura Luti, Federica Galaverna, Raffaella De Santis, Letizia Brescia, Linda Meneghello, Angelamaria Petrone, Nagua Giurici, Fabian Schumacher, Federico Mercolini

## Abstract

Vaccines represent the best tool to prevent the severity course and fatal consequences of pandemic by new Coronavirus 2019 infection (SARS-CoV-2). Considering the limited data on vaccination of pediatric oncohematological patients, we develop a Consensus document to support the Italian pediatric hematological oncological (AIEOP) centers in a scientifically correct communication with families and patients and to promote vaccination. The topics of the Consensus were: SARS-CoV-2 infection and disease (COVID-19) in the pediatric subjects; COVID-19 vaccines (type, schedule); which and when to vaccinate; contraindications and risk of serious adverse events; rare adverse events; third dose and vaccination after COVID-19; and other general prevention measures. Using the Delphi methodology for Consensus, 21 statements and their corresponding rationale were elaborated and discussed with the representatives of 31 centers, followed by voting. AIEOP Centers showed an overall agreement on all the statements that were therefore approved.

This consensus document highlights that children and adolescents affected by hematological and oncological diseases are a fragile category. Vaccination plays an important role to prevent COVID-19, to permit the regular administration of chemotherapy or other treatments, to perform control visits and hospital admissions, and to prevent treatment delays.

## Introduction

The persistence of the pandemic for the SARS-CoV-2 infection [1], the increase of infection rate in the pediatric population [2], the possibility of preventing infection through mRNA vaccines [3-5], and the limited evidence for pediatric oncohematological patients, induced the Infectious Working Group (IWG) of the Italian Association of Pediatric Hematology Oncology (AIEOP) to develop a Consensus document on COVID-19 vaccination. Currently, BNT162b2 vaccine (Pfizer/BioNTech-Comirnaty) and mRNA-1273 vaccine (Moderna-Spikevax) are authorized for the adolescent age (12-17 years) whereas BNT162b2 vaccine (Pfizer/ BioNTech-Comirnaty) is also authorized for 5-11 year-old children [6]. The aim of this initiative was to support AIEOP Centers for a scientifically correct communication with families and patients and to encourage sharing indications and recommendations for COVID-19 vaccination among pediatric hematology oncology centers.

## Methodology

A modified Delphi consensus was conducted between November and December 2021 by IGW-AIEOP following a methodology already published [7] that was based on the following steps: 1) establishment of scientific committee among members of IWG-AIEOP (SC, FM, FC, MC, AC, PM, MP, KP, FS) to define the topics; 2) assessment of the most recent relevant literature and the indications of the competent authorities (Food and Drug Administration (FDA), Advisory Committee on Immunization Practices (ACIP), European Medicine Agency (EMA), Italian Drug Agency (AIFA) on COVID-19 vaccination for pediatric age; 3) formulate summary statements and the corresponding scientific rationale among the scientific committee members followed by discussion of comments and criticisms; 4) final approval of statements by the scientific committee; 5) discussion and vote of statements in plenary meeting with the representatives of 31 AIEOP participating centers; 6) approval of the results of plenary session by scientific committee or, if needed, make a second-round of evaluation for those statements on which no consensus was achieved in the first round; 7) formulation of the consensus-based statements.

The degree of agreement or disagreement for each statement was evaluated using a Lykert scale from 1 to 5: 1 = strong disagreement, 2 = disagreement, 3 = mild agreement, 4 = agreement, 5 = strong agreement.

The results of voting were then aggregated according to this scheme: the sum of the scores of grade 1-2 were considered an expression of disagreement on the statement, while the sum of the scores of grades 3-5 were considered an expression of agreement on the statement.

The consensus for each sentence was considered reached when the sum of the scores 3-5 was ≥ 66% of the voters, while the non-consensus on the sentence was defined by a sum of the scores 1-2 > 66% of the voters. The strength of consent or non-consent was assessed by describing the frequencies of the individual scores for each sentence, being 5 the maximum score for consent and 1 for non-consent.

The scientific committee defined the following topics as objects of the Consensus: 1) SARS-CoV-2 and COVID-19 infection in the pediatric cancer patients; 2) type of vaccine; 3) vaccination schedule; 4) which oncohaematological patients to vaccinate; 5) when to vaccinate; 6) contraindications to vaccination and risk of serious adverse events; 7) rare adverse events of vaccination; 8) third dose and vaccination after COVID-19; 9) other general prevention measures. The rationale and the corresponding statements for each topic are presented in the below paragraphs, while the results of the vote are showed in table 1.

**Table 1.** List of statements and the aggregate score of the vote of the Centers

|  |  | Score after vote, % |  |  |  |  |  |
| --- | --- | --- | --- | --- | --- | --- | --- |
| N° | Statement | 1 | 2 | 3 | 4 | 5 | Tot. |
| 1 | *Pediatric oncohematological patients under active oncological treatment are considered frail subjects with a greater risk of severe COVID-19, critical COVID-19 or death from COVID-19 than the general immunocompetent pediatric population* | 0 | 0 | 6.5 | 19.4 | 74.2 | 100 |
| 2 | *In light of the individual risk and the emergence of highly contagious variants, pediatric oncohematological patients have a priority indication for COVID-19 vaccination* | 0 | 0 | 0 | 25.8 | 74.2 | 100 |
| 3 | *In the pediatric (5-11 years) and adolescent (12-17 years) populations, COVID-19 mRNA vaccines have a favorable ratio in terms of efficacy and adverse events* | 0 | 0 | 6.5 | 38.7 | 54.8 | 100 |
| 4 | *Vaccination with COVID-19 mRNA vaccine is indicated in pediatric oncohematological patients aged > 5 years* |  |  | 3.2 | 22.6 | 74.2 | 100 |
| 5 | *The primary vaccination course for immunocompromised pediatric and adolescent patients must comply with the vaccination schedule (vaccine dose, number of doses, interval between doses) recommended for the general pediatric and adolescent population.* | 0 | 0 | 3.2 | 12.9 | 83.9 | 100 |
| 6 | *As for other vaccines based on recombinant viral subunits, toxoid, viral protein components or conjugated bacterial polysaccharides, the immunocompromission condition is not a contraindication to receive a COVID-19 mRNA vaccine.* | 0 | 0 | 6.5 | 19.4 | 74.2 | 100.0 |
| 7 | *7) vaccination against COVID-19 is indicated:*<br>*a) in patients with haematological cancer (leukemia, lymphoma, myelodysplasia)*<br>*b) in patients with solid tumor or brain tumor*<br>*c) in patients undergoing autologous or allogeneic haematopoietic stem cell transplantation*<br>*d) in patients with primary immunodeficiency or with dysregulation of the immune system requiring chemotherapy or immunosuppression (eg. histiocytosis)*<br>*e) in patients undergoing therapy with biological drugs with an immunosuppressive effect*<br>*f) in patients undergoing cell therapy with chimeric receptor expression on T lymphocytes (CAR-T)*<br>*g) in patients with severe cytopenia of autoimmune origin or bone marrow failure*<br>*h) in patients with splenectomy or functional asplenia* | 0 | 0 | 3.2 | 25.8 | 71.0 | 100 |
| 8 | *Vaccination against COVID-19 should be performed in the clinically stable, non-febrile patient not undergoing steroid therapy > 0.5 mg/kg/day.* | 0 | 0 | 10 | 46 | 43 | 100 |
| 9 | *In patients receiving active chemotherapy, respect the following time intervals before vaccination:*<br>*a) from the last administration of anthracyclines: 7 days*<br>*b) since the last administration of high-dose cytarabine (cumulative dose > 6000 mg / m<sup>2</sup>): 4 weeks*<br>*c) from the administration of anti-PD-1 / PD-1L: 6-8 weeks* | 0 | 0 | 9.7 | 35.5 | 51.6 | 100 |
| 10 | *In patients undergoing cellular therapy (transplant or CAR-T), vaccinate at least 3 months after the procedure* | 0 | 0 | 3.2 | 35.5 | 61.3 | 100 |
| 11 | *In patients undergoing therapy with anti-CD20 monoclonal antibody (rituximab) or therapy with other monoclonals that cause severe and persistent B-lymphocytopenia, vaccinate at least 6 months after the last treatment* | 0 | 0 | 19.4 | 16.1 | 64.5 | 100 |
| 12 | *The history of previous PEG anaphylaxis or anaphylaxis after a first dose of mRNA vaccine is a contraindication respectively to the mRNA vaccine or to the second dose with mRNA vaccine* | 0 | 3.2 | 3.2 | 25.8 | 67.7 | 100.0 |
| 13 | *A previous allergic reaction to PEG-Asparaginase does not contraindicate the administration of the COVID-19 mRNA vaccine* | 0 | 3.2 | 16.1 | 32.3 | 48.4 | 100 |
| 14 | *A previous allergic reaction to other vaccines, allergy to foods, drugs, inhalants, insects, asthma are not contraindications to COVID-19 vaccination with mRNA vaccine* | 0 | 0 | 0 | 43.3 | 56.7 | 100.0 |
| 15 | *Vaccination of subjects considered to be at greater risk of anaphylaxis due to a personal history of severe allergy or comorbidities (mastocytosis, uncontrolled asthma) must be managed in collaboration with the allergist.* | 0 | 0 | 3.2 | 32.3 | 64.5 | 100 |
| 16 | *COVID-19 vaccination with a mRNA vaccine should not be performed in patients with clinical signs of myocarditis, pericarditis, rheumatic heart disease or myocardial contractility deficiency, but postponed until their resolution* | 0 | 0 | 3.2 | 16.1 | 80.6 | 100 |
| 17 | *The potential risk of immune thrombocytopenia does not represent a reason to contraindicate COVID-19 vaccination* | 0 | 0 | 0 | 29 | 71 | 100 |
| 18 | *Pediatric or adolescent oncohematological patients are advised to receive a third dose of mRNA vaccine, or in the case of a previous SARS-Cov-2 infection, complete vaccination with 2 dose schedule* | 0 | 0 | 0 | 12.9 | 87.1 | 100 |
| 19 | *The third dose is indicated at least 4 weeks after the second dose for patient groups in which a reduced response is expected (e.g. CAR-T, lymphoma, leukemia) or documented (antibody control), or 5-6 months after second dose to* *prevent antibody decline or according to the most recent provisions of the regulatory authorities; in the event of a previous SARS-CoV-2 infection, complete vaccination at 2 doses is indicated starting from 3 months after infection.* | 0 | 0 | 3.2 | 22.6 | 74.2 | 100 |
| 20 | *COVID-19 vaccination of family members and close cohabitants is recommended, including the third dose, in accordance with the indications of the regulatory authorities for the various age groups* | 0 | 0 | 0 | 6.5 | 93.5 | 100 |
| 21 | *Personal protective equipment such as filter masks, hand hygiene and physical or social distancing remain the cornerstones of preventing SARS-CoV-2 infection in pediatric oncohematological patients* | 0 | 0 | 0 | 6.5 | 93.5 | 100 |

### SARS-CoV-2 and COVID-19 infection in the pediatric cancer patients

The SARS-CoV-2 infection developed in China from December 2019 and rapidly spread throughout the world, so that on 11 March 2020 the World Health Organization declared a pandemic status, which is still in force today.

The SARS-CoV-2 virus has proved highly contagious, spreading by respiratory tract (droplets, aerosols) or by direct contact of the mucous membranes with infected biological fluids.

Although the infection can occur asymptomatically or paucisymptomatically up to 80% of cases, in the remaining cases it can be severe or very severe. The symptomatic form of SARS-CoV-2 infection, called novel Coronavirus-2019 disease (COVID-19) has caused, at the time of writing this paper, more than 5 million deaths [8]. The main risk factor for a severe form of COVID-19 or for COVID-19-related mortality is older age. Other risk factors are the presence of obesity, cardiovascular diseases, chronic lung and kidney diseases, as well as the presence of tumors in an active phase of treatment. In the pediatric population, susceptibility to SARS-CoV-2 infection was lower, with a lower incidence of severe or critical forms and a lower mortality (0.01%-0.7%)[9,10]. Nevertheless, pediatric oncohematological patients may represent a population at greater risk of morbidity and mortality from COVID-19 due to the greater susceptibility of immunosuppressed patients to respiratory infections, the impairment of innate immunity during oncological treatments with chemotherapy and/or radiotherapy, the immunosuppression associated with stem cell transplant procedures and the need for frequent access to hospital facilities for visits and treatments.

In the centers of the AIEOP network, the necessary measures to keep free the wards from COVID-19 have been put in place since the end of February 2020, such as triage for fever, respiratory infection, and contact with COVID-19 subjects, separate paths for COVID-19 positive patients, adequate information to parents, caregivers and patients, implementation of hygiene measures (washing/ cleaning hands), use of personal protective equipment (mask, visor, gloves) by health operators, screening of all patients before admissions for chemotherapy, anesthesia or sedation for surgery or invasive or diagnostic procedures by nasopharyngeal molecular swab for SARS-CoV-2 [11]. This allowed to limit the incidence of COVID-19 in AIEOP centers. Moreover, the clinical course of pediatric oncohematological SARS-CoV-2-positive patients was mostly favorable, with only 3% of them requiring hospitalization in intensive care and no COVID-19-related death observed during the first pandemic wave (March-May 2020) and the second and third wave (October 2020-May 2021)[12,13].

Although the experience of the AIEOP centers has so far been comforting, global data showed that COVID-19 can have a serious course in pediatric patients suffering from oncohematological disease. In the period 15 April 2020-1 February 2021, 1520 pediatric oncohematological patients with SARS-CoV-2/COVID-19 were registered in the Global COVID-19 Registry, of which 1319 (87%) had a minimum follow-up of 30 days; of them, 30.5% were asymptomatic, 67.4% were hospitalized, and 55.8% changed their oncological plan of treatment due to infection. The severe or very severe forms of COVID-19 were 19.9% while the COVID-19-realted mortality was 3.8% [14]. In multivariate analysis, the risk factors for COVID-19 severe or very severe were the country’s low or medium-low per capita income (according to World Bank data), age 15-18 years, lymphopenia <300/mmc, neutropenia <500/mmc, and the need for intensive care. Factors associated with the modification of chemotherapy or oncological plan due to SARS-CoV-2 infection were the upper-middle income per capita of the country, the diagnosis of hematological cancer (compared with the diagnosis of solid tumor), the presence of one or more symptoms at the time of infection, the diagnosis of COVID-19, and the presence of one or more comorbidities. In this study, the average income per capita, in addition to clinical parameters, influenced significantly the impact of the pandemic on clinical outcomes; in fact, a 5.8 time-higher relative risk of severe or critical COVID-19 was observed in low-or lower-middle-income countries, as opposed to high-income ones. This is consistent with the fact that the country average-per-capita-income affects the country general health care organization, the level of infrastructure to access the health care services, and the availability of hospitals and intensive care units (number of beds per thousand inhabitants). Therefore, the risk of a serious or fatal course of SARS-CoV-2 infection in the pediatric oncohematological patient population is variable as it depends on the extent of the immunosuppression to which the patient is subjected, on the therapeutic and intervention capacity of the health system in which the patient is treated and on the contagiousness and virulence of the SARS-CoV-2 strain predominant at a given time or geographical area. In this regard, the Delta variant of SARS-CoV-2, predominant in Italy and Europe during the second part of 2021, has been shown to be more contagious in children than the wild strain of SARS-CoV-2 [15]. Moreover, a new variant of concern has been identified in South Africa 26 November 2021, named as Omicron [16], that is rapidly spreading also in Europe [17] and creating worldwide concerns owing to its higher transmissibility and partial resistance to immunity induced by vaccines and monoclonal antibody-based therapies [18-20].

#### Statements

1 *Pediatric oncohematological patients under active oncological treatment are considered frail subjects with a greater risk of severe COVID-19, critical COVID-19 or death from COVID-19 than the general immunocompetent pediatric population*.
2 *In light of the individual risk and the emergence of highly contagious variants, pediatric oncohematological patients have a priority indication for COVID-19 vaccination*

### Type of vaccine

The vaccines in Europe approved by EMA are 5: 2 are non-replicative adenoviral vector vaccines (AstraZeneca/Vaxzevria, Janssen/Johnson & Johnson), 2 are mRNA vaccines (Pfizer/BioNTech-Comirnaty; Moderna-Spikevax) and one is a protein-based vaccine (Novavax-Nuvaxovid, also known as NVX-CoV2373) [21]. Of these, only mRNA vaccines are indicated for the age 5-17 years because data of efficacy and safety are available [22]. In a randomized placebo-controlled study of 2260 subjects aged 12-15 years, 2 doses of BNT162b2 vaccine (Pfizer/BioNTech-Comirnaty), administered with an interval of 21 days, showed 100% efficacy against COVID-19, while the adverse events was mild to moderate, limited to the first 24-48 hours post-1st and 2nd dose (injection site pain 79-86%, fatigue 60-66%, headache 55-65%); no serious adverse events were recorded. The antibody response (neutralized antibodies) was also superior to that of a control population aged 16-25 years [3].

In a randomized placebo-controlled study of 3732 subjects aged 12-17 years, 2 doses of mRNA-1273 vaccine (Moderna-Spikevax), administered with an interval of 28 days, showed an efficacy against COVID-19 of 93.3%. Adverse events were mostly mild and moderate post-1st and 2nd dose (injection site pain 93% -92%, headache 44% -70%, fatigue 48% -68%), limited to the first 48 hours post-injection and no serious adverse events were recorded. The neutralizing antibody titers were comparable to that of a control group aged 18-25 years. The ability of the vaccine to prevent asymptomatic SARS-CoV-2 infection was also evaluated in this study, which was 59.5% 14 days after the first dose and 39.2% 14 days after the second dose [4].

A higher incidence of myocarditis has been reported with the use of mRNA vaccines in adolescent and young adults, in particular with the mRNA-1273 (Moderna-Spikevax) vaccine for which the use of the BNT162b2 (Pfizer / BioNTech-Comirnaty) vaccine is recommended for subjects aged <18 years [23,24].

Recently, FDA and EMA issued the authorization for the emergency use of the BNT162b mRNA vaccine (Pfizer/BioNTech-Comirnaty) for the age group 5-11 years based on a randomized study enrolling 4,700 subjects [5,25]. The dose of messenger mRNA used in this pediatric study was 10 micrograms instead of the 30 micrograms used in the studies with subjects > 12 years of age. The analysis showed that, 7 days after the second dose, the effectiveness of the vaccine was 90.7%. Safety data on more than 4,600 subjects, including 1,444 vaccinated for at least 2 months after the second dose, showed that side effects were generally mild to moderate in severity, generally limited to two days following vaccination and more frequent after the second dose than the first. Commonly reported side effects were injection site pain (arm pain), redness and swelling, fatigue, headache, muscle and or joint pain, chills, fever, swollen lymph nodes, nausea and decreased appetite. In a subgroup of 264 study subjects, the immunogenicity of the vaccine was assessed and it resulted comparable with that of a control group of 253 subjects, aged 16-25 years, who had received two doses of the vaccine in a previous study.

Unlike the general pediatric population, the experience of COVID-19 vaccination in pediatric cancer patients is limited. In a French study, 11 adolescents and young adults aged 16 to 21 years (5 of 16 years, 3 of 17 years, 2 of 18 years and 1 of 21 years), suffering from solid cancer undergoing active treatment or within 6 months of stopping chemotherapy, they were vaccinated with 2 doses of BNT162b mRNA vaccine (Pfizer/BioNTech-Comirnaty) obtaining a neutralizing antibody response in 9 out of 10 patients (1 was already positive in serology before the first dose) [26].

#### Statements

3 *In the pediatric (5-11 years) and adolescent (12-17 years) populations, COVID-19 mRNA vaccines have a favorable ratio in terms of efficacy and adverse events*.
4 *Vaccination with COVID-19 mRNA vaccine is indicated in pediatric oncohematological patients aged > 5 years*.

### Vaccination schedule

There are no specific data about the vaccination schedule for immunocompromised patients aged > 5 years; therefore, pending specific studies, the indication for these patients is to use the same vaccination schedule approved for the immunocompetent population aged > 5 years.

#### Statements

5 *The primary vaccination course for immunocompromised pediatric and adolescent patients must comply with the vaccination schedule (vaccine dose, number of doses, interval between doses) recommended for the general pediatric and adolescent population*.

### Which oncohaematological patients to vaccinate

The immunosuppressed condition does not represent a contraindication to vaccination, with the exception of vaccines based on live attenuated viruses and the tuberculous vaccine, but a further motivation for vaccination to protect the more fragile patients [27]. The patient’s immunocompromission may cause a reduced response and, therefore, a lower efficacy of vaccine, but, the lower immune response can determine a better tolerability due to a lower incidence of local or systemic side effects.

In order to identify the frail patients who deserve the vaccination because of oncological treatment, the expert group referred to the list of drugs and clinical conditions associated with immunocompromission provided by AIFA and CDC [28, 29].

#### Statements

6 *As for other vaccines based on recombinant viral subunits, toxoid, viral protein components or conjugated bacterial polysaccharides, the immunocompromission condition is not a contraindication to receive a COVID-19 mRNA vaccine*.
7 *Vaccination against COVID-19 is indicated:*

a. *in patients with haematological cancer (leukemia, lymphoma, myelodysplasia)*
b. *in patients with solid tumor or brain tumor*
c. *in patients undergoing autologous or allogeneic haematopoietic stem cell transplantation*
d. *in patients with primary immunodeficiency or with dysregulation of the immune system requiring chemotherapy or immunosuppression (eg. histiocytosis)*
e. *in patients undergoing therapy with biological drugs with an immunosuppressive effect*
f. *in patients undergoing cell therapy with chimeric receptor expression on T lymphocytes (CAR-T)*
g. *in patients with severe cytopenia of autoimmune origin or bone marrow failure*
h. *in patients with splenectomy or functional asplenia*

### When to vaccinate

Vaccination against COVID-19 is to be considered a priority, also compared to other vaccinations, given the persistence of the emergency pandemic state, the continuous viral circulation with resurgence, especially in the autumn and winter season, the emergence of more contagious variants such as the variant Delta and the Omicron variants, and, for the pediatric age, the lack of preventive drugs to manage the pre-or-post-exposure to the virus or in the paucisymptomatic phase (e.g. monoclonal anti-spike casirivimab/imdevimab for age < 12 years or sotrovimab or the new antiviral molnupiravir for age <18 years). The choice of when to vaccinate a patient under treatment is under discussion as specific studies have not been conducted. Therefore, the decision of vaccination must be personalized on the basis of the intensity and the type of treatment in progress. This is for two reasons: 1) to avoid the overlap of rare, unexpected adverse events of the vaccine with acute organ toxicity due to chemotherapy treatment or treatment with biological drugs; 2) allow the immune system to recover its vaccine response capacity. In pediatric cancer patients, the immune response to vaccines has been observed during chemotherapy in subjects with a lymphocyte value > 700 / mmc, with underlying disease in remission and in the absence of steroid therapy, intensive chemotherapy, and infections [30].

In adolescents and young adults (16-30 years), post-vaccination COVID-19 myocarditis has been reported, therefore it is advisable to distance vaccination from the administration of drugs associated with a risk of myocarditis and pericarditis such as anthracyclines, high doses of cytarabine and anti-PD-1 drugs. Anthracyclines can cause acute cardiotoxicity and pericarditis within one week of administration [31]. Cytarabine administered in high doses (> 6000 mg/m2) can cause cardiotoxicity and pericarditis up to 4 weeks after administration as part of a delayed type IV hypersensitivity reaction [32,33]. Anti-PD-1 drugs, immune checkpoint inhibitors, can be associated with the onset of pericarditis up to 6-8 weeks after their administration [34,35]. In subjects undergoing therapy with B-lymphocyte-depleting drugs, particularly rituximab, the almost complete absence of antibody vaccine response was observed within 6 months of the last administration [36]. In adult subjects undergoing CAR-T therapy, which induces a prolonged hypogammaglobulinaemia and marked B lymphopenia, vaccination performed at a median distance of 401 days (range, 113-819) from treatment showed a response of 21% after the first dose and 30% after the second dose [37]. In subjects undergoing allogeneic transplant, antibody and cellular vaccine responses (particularly in subjects with B-cell aplasia) were instead observed in 75% of patients at a median distance of 32 months (range 3-263) from transplantation [38]. The interval between cell therapy (CAR-T or transplant) and vaccination in the various studies is 3 months as this is considered the minimum interval to observe a recovery of the specific immune response.

#### Statements

8 *Vaccination against COVID-19 should be performed in the clinically stable, non-febrile patient not undergoing steroid therapy > 0*.*5 mg/kg/day*.
9 *In patients receiving active chemotherapy, respect the following time intervals before vaccination:*

a. *from the last administration of anthracyclines: 7 days*
b. *since the last administration of high-dose cytarabine (cumulative dose> 6000 mg / m2): 4 weeks*
c. *from the administration of anti-PD-1 / PD-1L: 6-8 weeks*

10 *In patients undergoing cellular therapy (transplant or CAR-T), vaccinate at least 3 months after the procedure*
11 *In patients undergoing therapy with anti-CD20 monoclonal antibody (rituximab) or therapy with other monoclonals that cause severe and persistent B-lymphocytopenia, vaccinate at least 6 months after the last treatment*

### Contraindications to vaccination and risk of serious adverse events

mRNA vaccines are formulated as lipid nanoparticles coated with polyethylene glycol (PEG), a substance used in many pharmaceutical or cosmetic products. A previously documented PEG allergy is a contraindication to the mRNA vaccine. Another contraindication to the second dose of mRNA vaccine is the anaphylactic reaction to the first dose, which incidence is approximately 4.7 cases per million doses administered [39]. In these cases, vaccination can be performed with a vaccine different from the mRNA vaccine, if authorized for age.

Non-anaphylactic allergic reactions to a first dose of vaccine are not a contraindication to administration of the second dose. In this case, the decision to proceed will be evaluated after consulting the allergist specialist and it is advisable to be made under medical observation. On the other hand, previous allergic reactions to other vaccines, allergies to drugs, foods, inhalants, insect venom, asthma are not contraindications to COVID-19 vaccination and must be managed with a more prolonged observation (30-60 minutes instead of the standard 15 minutes). In case of subjects with a personal history of severe allergies to multiple drugs or with comorbidities at risk of anaphylaxis (mastocytosis, uncontrolled asthma), vaccination must be managed in collaboration with the allergist [40].

Individuals with an allergic reaction to PEG-asparaginase have no absolute contraindication to COVID-19 vaccination. In fact, it is possible that the allergy is due to the L-asparaginase instead of the PEG component. The type of PEG used in mRNA vaccines is different from that used in other drugs and preparations for which cross-reactivity is uncertain. In 2 studies, in which 32 and 19 patients, with a history of PEG-L-Asparaginase allergy, were vaccinated with 2 doses of BNT162b vaccines (Pfizer-BioNTech), no patients developed any signs or symptoms of allergy [41,42]. In these cases, it is recommended to proceed after consulting the allergist and carry out a more prolonged observation of the subject at the vaccination site (30-60 minutes instead of the standard 15 minutes).

#### Statements

12 *The history of previous PEG anaphylaxis or anaphylaxis after a first dose of mRNA vaccine is a contraindication respectively to the mRNA vaccine or to the second dose with mRNA vaccine*.
13 *A previous allergic reaction to PEG-Asparaginase does not contraindicate the administration of the COVID-19 mRNA vaccine*.
14 *A previous allergic reaction to other vaccines, allergy to foods, drugs, inhalants, insects, asthma are not contraindications to COVID-19 vaccination with mRNA vaccine*.
15 *Vaccination of subjects considered to be at greater risk of anaphylaxis due to a personal history of severe allergy or comorbidities (for example mastocytosis, uncontrolled asthma) must be managed in collaboration with the allergist*.

### Rare adverse events

Although the safety and tolerability profile of mRNA vaccines is favorable, myocarditis and pericarditis have been reported as a rare complication, mainly in young male subjects less than 30 years of age who received the mRNA vaccines [43]. In Israel, where the population received the BNT162b Pfizer-BioNTech mRNA vaccine the incidence of myocarditis was 1:26,000 in males and 1:228,000 in females. Compared to the unvaccinated population, the highest incidence was observed for the age group of 16-25 years [44]. In two recent European epidemiological studies, one in France and one conducted in the Nordic countries, it showed that myocarditis and pericarditis are “very rare “with an incidence of approximately 1:10,000 vaccinated. Complication occurs a few days to 14 days after administration and is more common in young males [45].

Furthermore, the incidence of myocarditis/pericarditis is higher after mRNA-1273 vaccine (Moderna-Spikevax), and is 1.3: 10,000 cases in the French study (age group 12-29 years), and in the Nordic study (age group 16-24 years), 1.9: 10,000 cases compared to unvaccinated; while in the mRNA vaccine BNT162b (Pfizer-BioNTech) the incidence of myocarditis/pericarditis was 0.26: 10,000 in the French study and 0.57 in the Nordic study. Considering these data, it is advisable to prefer the BNT162b mRNA vaccine (Pfizer-BioNTech) in subjects < 18 years of age [46,47]. The clinical presentation of myocarditis after vaccination was generally mild with response to conservative or symptomatic treatment. [47].

Unlike adenoviral vector vaccines (AstraZeneca/VaxZevria and Janssen/COVID-19), no serious complication of vaccine-induced thrombosis/thrombocytopenia (VIIT) was reported in mRNA vaccines [48]. On the other hand, cases of immune thrombocytopenia appearing in the first 2 weeks after vaccination (usually the first dose) have been reported: 17 cases in the United States in the period January-February 2021 in which 20 million people were vaccinated [49]. Thrombocytopenia immune is also reported for other vaccinations, for example 1:40,000 children vaccinated for measles-rubella-mumps, but the rarity of the recorded events does not allow the distinction between the event induced by vaccination (causal relationship) and event coinciding with vaccination (random relationship), as immune thrombocytopenia occurs in the pediatric or adult population even beyond any temporal correlation with vaccinations. These aspects underline the importance of population surveillance to identify rare adverse events, with an incidence of less than 1:100,000-1:1,000,000, difficult to detect in authorization studies where a more limited number of subjects are evaluated.

#### Statements

16 *COVID-19 vaccination with a mRNA vaccine should not be performed in patients with clinical signs of myocarditis, pericarditis, rheumatic heart disease or myocardial contractility deficiency, but postponed until their resolution*.
17 *The potential risk of immune thrombocytopenia does not represent a reason to contraindicate COVID-19 vaccination*.

### Third dose and vaccination after COVID-19

In oncohaematological and immunosuppressed patients, a third dose of vaccine is required once the vaccination cycle is completed with 2 doses of mRNA vaccine. The third dose of the vaccine is defined as 1) “extended dose” or additional dose, or 2) “booster dose” [50-53].

The extended dose is an additional dose administered at least 4 weeks after the second vaccination dose in patients belonging to those fragile categories in which a reduced antibody response is expected. The dose of mRNA vaccine is identical to that used for the primary vaccine cycle, i.e. 30 ug for BNT162b (Pfizer-BioNTech) and 100 ug for mRNA-1273 (Moderna-Spikevax). In oncohaematological patients under treatment, in transplant patients, in immunosuppressed or immunodeficient patients, there is therefore an indication to receive a third dose at least 28 days after the completion of the vaccination cycle [54].

The booster dose is instead a third dose administered from 5-6 months after the completion of the vaccination cycle in order to counteract the decline, over time, of the antibody response in subjects with normal immune systems and aged > 18 years. The dose of mRNA vaccine is identical to that used for the primary vaccination cycle for BNT162b (Pfizer-BioNTech), 30 ug, while it is reduced to 50 ug for mRNA-1273 (Moderna-Spikevax). The booster dose is a priority particularly in subjects belonging to categories at high risk of exposure to the infection, such as healthcare personnel, but it is to be recommended, even to parents and close family members of frail patients [55,56].

Complete vaccination (2 doses), instead of a single dose as in the immunocompetent, is indicated in oncohematological and immunosuppressed patients who have contracted SARS-CoV-2 infection both asymptomatically and symptomatically (COVID-19) from the 3rd month after the infection [57].

#### Statements

18 *Pediatric or adolescent oncohematological patients are advised to receive a third dose of mRNA vaccine, or in the case of a previous SARS-Cov-2 infection, complete vaccination with 2 dose schedule*.
19 *The third dose is indicated at least 4 weeks after the second dose for patient groups in which a reduced response is expected (e*.*g. CAR-T, lymphoma, leukemia) or documented (antibody control), or 5-6 months after second dose to prevent antibody decline or according to the most recent provisions of the regulatory authorities; in the event of a previous SARS-CoV-2 infection, complete vaccination at 2 doses is indicated starting from 3 months after infection*.

### Other general prevention measures

Vaccination does not give the individual absolute and lasting protection over time. Furthermore, there are population groups in which the virus circulates widely (non vax, no mask, pediatric population). Therefore the risk of contagion remains high especially in the months of the year when the circulation of the virus increases favored by climatic reasons or where there are concentrations of people.

Pending the extension of vaccination to the entire pediatric population, vaccination of parents and close family members and the maintenance of individual and social protection measures such as the use of a mask, hand hygiene and physical or social distancing are recommended. In this context, the annual flu vaccination for the patient and family members remains a recommended preventive measure [58].

#### Statements

20 *COVID-19 vaccination of family members and close cohabitants is recommended, including the third dose, in accordance with the indications of the regulatory authorities for the various age groups*.
21 *Personal protective equipment such as filter masks, hand hygiene and physical or social distancing remain the cornerstones of preventing SARS-CoV-2 infection in pediatric oncohematological patients*.

### Final comment

The vote of AIEOP Centers showed an overall agreement on all the statements (score from 3 to 5> 66% of the voters for all 21 statements). No center showed strong disagreement in any of the proposed statements which are therefore approved.

Only one center of the 31 voters expressed disagreement with statements 12 and 13 relating to the indication of vaccination in subjects allergic to PEG or with a previous allergic reaction to PEG-Asparaginase. For these 2 statements, the voting of the other centers was also more distributed. This demonstrates that the management of the patient at risk of severe allergic reaction requires a careful evaluation of the risks and benefits personalized on the patient and in a multidisciplinary context.

This document highlights how the community of Italian oncohematological pediatricians recognizes that patients suffering from hematological and oncological diseases, represent a wide category of fragile children in which vaccination plays an important role to prevent SAS-CoV-2 infection and COVID-19, to allow the regular chemotherapy or immunosuppressive treatment and check-ups or hospital admissions and to prevent treatment delays.

## Data Availability

All data produced in the present work are contained in the manuscrip

